# *APOE*ε4 carriership associates with microglial activation independently of Aβ plaques and tau tangles

**DOI:** 10.1101/2022.07.12.22277556

**Authors:** João Pedro Ferrari-Souza, Firoza Z. Lussier, Douglas T. Leffa, Joseph Therriault, Cécile Tissot, Bruna Bellaver, Pâmela C. Lukasewicz Ferreira, Maura Malpetti, Yi-Ting Wang, Guilherme Povala, Andréa L. Benedet, Nicholas J. Ashton, Mira Chamoun, Stijn Servaes, Gleb Bezgin, Min Su Kang, Jenna Stevenson, Nesrine Rahmouni, Vanessa Pallen, Nina Margherita Poltronetti, John T. O’Brien, James B. Rowe, Ann D. Cohen, Oscar L. Lopez, Dana L. Tudorascu, Thomas K. Karikari, William E. Klunk, Victor L. Villemagne, Jean-Paul Soucy, Serge Gauthier, Diogo O. Souza, Henrik Zetterberg, Kaj Blennow, Eduardo R. Zimmer, Pedro Rosa-Neto, Tharick A. Pascoal

## Abstract

Microglial activation is an early phenomenon in Alzheimer’s disease (AD) that may occur prior to and independently of amyloid-β (Aβ) aggregation. Recent studies in transgenic animal models suggest that the apolipoprotein E ε4 (*APOE*ε4) allele may be a culprit of early microglial activation in AD. However, it is unclear whether the *APOE*ε4 genotype is associated with microglial reactivity in the living human brain. Here, we tested whether *APOE*ε4 carriership is associated with microglial activation in individuals across the aging and AD spectrum. We studied 118 individuals who had positron emission tomography (PET) for Aβ ([^18^F]AZD4694), tau ([^18^F]MK6240), and microglial activation ([^11^C]PBR28), as well as clinical, genetic, and magnetic resonance imaging data. We found that *APOE*ε4 carriership was associated with increased microglial activation mainly in early Braak-staging regions within the medial temporal cortex, and this effect of *APOE*ε4 was independent of Aβ and tau deposition. Furthermore, microglial activation mediated the Aβ-independent effects of *APOE*ε4 on downstream tau accumulation, neurodegeneration, and clinical impairment. Interestingly, the physiological distribution of *APOE* mRNA expression, obtained from the Allen Human Atlas, predicted the patterns of *APOE*ε4-related microglial activation in our population, suggesting that the deleterious effects of *APOE*ε4 occur at the level of gene expression. These results support a model in which the *APOE*ε4 has Aβ-independent effects on AD pathogenesis by activating microglia in brain regions associated with early tau deposition. Our findings provide a rationale for the development of novel AD therapies targeting the interplay between ApoE and neuroinflammation.

## Introduction

Alzheimer’s disease (AD) is a multifactorial disorder neuropathologically characterized by amyloid-β (Aβ) plaques and tau neurofibrillary tangles *(1, 2)*. Among the multiple pathogenic processes involved in AD etiology, neuroinflammation - commonly associated with microglial reactivity - has been increasingly recognized *(3, 4)*. Microglial activation plays a key role in the accumulation of AD hallmark proteinopathies, rather than being merely an epiphenomenon of their deposition *(3, 4)*. Specifically, recent observations from animal and human studies suggest that microglial activation precedes and may drive tau spread over the neocortex following a Braak stage-like pattern *(5-7)*, from the medial temporal to association and primary sensory structures *(8-11)*. Such microglial activation is synaptotoxic, affects brain connectivity, and predicts clinical decline *(12, 13)*. Aβ pathology can trigger microglial activation in AD *(14-16)*, but Aβ plaques and activated microglia only partially overlap topographically in the human brain *(17-19)*, and microglial activation may occur prior to demonstrable Aβ deposition *(3)*.

The apolipoprotein E ε4 (*APOE*ε4) allele is a major genetic risk factor for sporadic AD *(20-23)*. The link between *APOE*ε4 and Aβ deposition is an important factor leading to AD progression *(24)*. However, recent animal studies suggest that the *APOE*ε4 genotype may also contribute to AD pathogenesis through Aβ-independent pathways by potentiating brain inflammation, tau accumulation, and neurodegeneration *(25, 26)*. Although robust experimental evidence indicates that the *APOE* genotype modulates microglial response in AD *(25, 27-30)*, it remains to be elucidated whether *APOE*ε4 carriership is associated with microglial activation in the AD human brain. The characterization of this association in living individuals is critical to confirm the Aβ-independent detrimental effects of *APOE*ε4 on microglia homeostasis and to support the development of novel therapeutic strategies.

Using complementary positron emission tomography (PET) radiotracers for the topographical quantification of microglial activation, Aβ, and tau accumulation across the brain, we investigated the association of *APOE*ε4 carriership, microglial activation, Aβ, and tau in a cohort of individuals across the aging and AD continuum. We hypothesized that *APOE*ε4 is associated with microglial activation independently of AD hallmark proteinopathies. We then tested whether microglial activation mediates the effects of *APOE*ε4 on tau accumulation, neurodegeneration, and clinical impairment. *Postmortem* data from the Allen Human Brain Atlas was used to test the link between regional levels of brain *APOE* gene expression and the distribution of microglial activation as a function of *APOE*ε4 carriership.

## Results

### Participants

We screened 606 people for the rs6971 polymorphism in the translocator protein (*TSPO*) gene. Out of the 314 high-affinity binders, we studied 118 individuals that were across the aging and AD spectrum (79 cognitively unimpaired [CU], 23 with mild cognitive impairment [MCI], and 16 with AD dementia) with [^18^F]AZD4694 Aβ PET, [^18^F]MK6240 tau PET, [^11^C]PBR28 microglial activation PET, and magnetic resonance imaging (MRI), as well as *APOE* genotyping (**Fig. S1**). Demographic characteristics of the population are reported in **Table 1**. Information regarding the prevalence of APOE genotypes in our sample is described in **Table S1**.

**Table 1.** Demographics and key characteristics of participants by clinical diagnosis.

|  | **CU** | **MCI** | **AD** |
| --- | --- | --- | --- |
| No. | 79 | 23 | 16 |
| Age, years | 72.3 (5.7) | 73.0 (5.3) | 70.1 (10.3) |
| Male, No. (%) | 17 (21.5) | 14 (60.9) | 9 (56.3) |
| Education, years | 15.2 (3.7) | 15.5 (2.9) | 13.6 (3.8) |
| *APOE*ε4 carrier, No. (%) | 23 (29.1) | 15 (65.2) | 7 (43.8) |
| MMSE score | 29.2 (1.0) | 28.2 (1.6) | 22.1 (5.9) |
| CDR-SB score | 0.1 (0.2) | 1.5 (0.8) | 5.3 (2.5) |
| Global [<sup>18</sup>F]AZD4694 SUVR | 1.52 (0.43) | 2.18 (0.58) | 2.42 (0.54) |
| Braak I-II [<sup>18</sup>F]MK6240 SUVR | 0.94 (0.19) | 1.38 (0.52) | 1.60 (0.39) |
| Braak III-IV [<sup>18</sup>F]MK6240 SUVR | 0.96 (0.10) | 1.28 (0.53) | 2.19 (1.13) |
| Braak V-VI [<sup>18</sup>F]MK6240 SUVR | 0.98 (0.09) | 1.16 (0.31) | 2.05 (1.22) |
| Hippocampal volume, cm<sup>3</sup> | 3.45 (0.34) | 3.18 (0.34) | 2.95 (0.55) |

Continuous variables are presented as mean (SD). Abbreviations: AD = Alzheimer's disease; *APOE*ε4 = Apolipoprotein E ε4; CDR-SB = clinical dementia rating scale sum of boxes; CU = cognitively unimpaired; MCI = mild cognitive impairment; MMSE = Mini-Mental State Examination; ROI = region of interest; SD = standard deviation; SUVR = standardized uptake value ratio.

### *APOE*ε4 associates with microglial activation in the medial temporal cortex

To test the association between *APOE*ε4 status and [^11^C]PBR28 microglial activation PET, we conducted linear regression analyses adjusting for age, sex, and clinical diagnosis. Voxel-wise regression analysis showed that *APOE*ε4 carriership was associated with higher [^11^C]PBR28 uptake mainly in medial temporal structures (transentorhinal, entorhinal, and hippocampal cortices), which are regions corresponding to early Braak stages (**Fig. 1A, B**). Regarding the spatial extent of the voxel-wise results, the association between *APOE*ε4 carriership and [^11^C]PBR28 (SUVR) was predominantly observed in areas corresponding to Braak I (affecting 94.8% of this region), followed by Braak II (47.6%), Braak III (16.7%), Braak IV (13.1%), Braak V (2.5%), and Braak VI regions (1.0%; **Fig. 1C**). Similarly, in terms of the magnitude of the associations (β estimate), the relationship between *APOE*ε4 and [^11^C]PBR28 uptake was progressively weaker from Braak I to VI, surviving Bonferroni correction for multiple comparisons only in Braak I and II regions (Braak I: β = 0.088, 95% confidence interval [CI] 0.044 to 0.133, *P* < 0.001; Braak II: β = 0.066, 95% CI 0.020 to 0.111, *P* = 0.005; **Fig. 1D**). Sensitivity analysis excluding individuals bearing the ε2 allele of the *APOE* gene showed similar findings (**Fig. S2**).

**Fig. 1.**
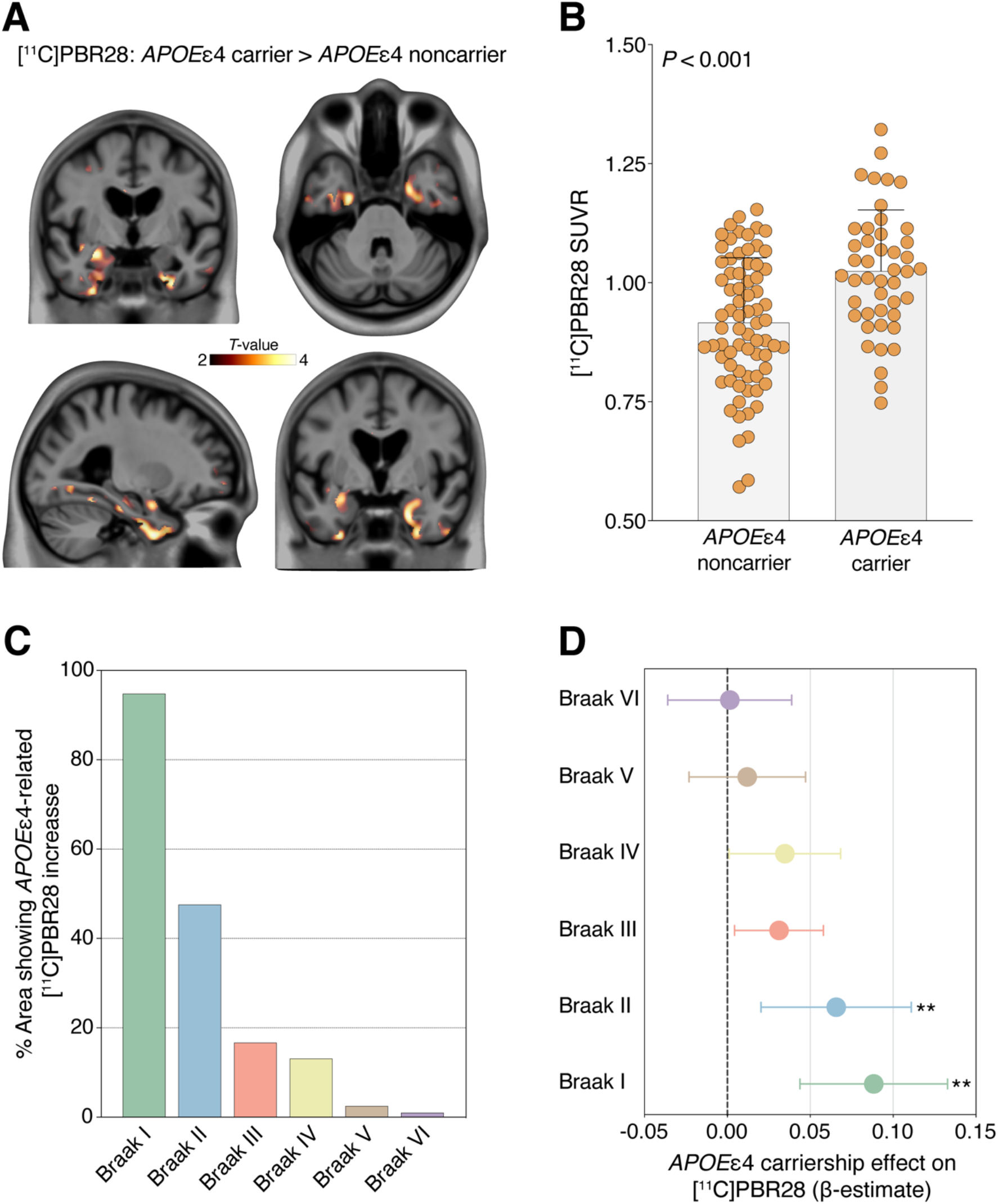
*APOE*ε4 carriership is associated with microglial activation in early Braak regions. **(A)** *T*-map shows the result of voxel-wise linear regression testing the association of *APOE*ε4 carriage status (noncarrier or carrier) with [^11^C]PBR28 SUVR accounting for age, sex, and clinical diagnosis (CU, MCI, or AD). Results survived random field theory correction for multiple comparisons at *P* < 0.05. (**B**) Bars show the mean and standard deviation of [^11^C]PBR28 SUVR in *APOE*ε4 noncarriers and carriers. Imaging biomarker values were extracted from the peak *T*-value cluster of the voxel-wise analysis (*T*-value ≥ 4.7). *P*-value indicates the result of regression analysis accounting for age, sex, and clinical diagnosis. (**C**) Bars represent the spatial extent of the *APOE*ε4*-*related microglia activation across Braak regions. Values were calculated by determining the percentage of voxels per Braak region having an association (*T*-value > 2) between *APOE*ε4 carriership and [^11^C]PBR28 in the voxel-wise analysis. (**D**) β estimates with 95% CI represent the strength of the regional association between *APOE*ε4 status and [^11^C]PBR28 SUVR across Braak regions from ROI-based linear regressions. Models were adjusted for age, sex, and clinical diagnosis. Estimates that survived Bonferroni correction at *P* < 0.05 are indicated with a double asterisk.

### *APOE*ε4 associates with microglial activation independently of Aβ and tau biomarkers

We investigated whether the association between *APOE*ε4 carriership and [^11^C]PBR28 uptake was independent of AD hallmark proteinopathies by correcting the regression models for global [^18^F]AZD4694 Aβ PET and local [^18^F]MK6240 tau PET, as well as age, sex, and clinical diagnosis. *APOE*ε4 carriership was associated with higher [^11^C]PBR28 uptake in Braak I-II regions independently of global Aβ and local tau PET (β = 0.055, 95% CI 0.010 to 0.100, *P* = 0.018; **Table 2**). In a subgroup of 51 participants (31 CU, 12 with MCI, and 8 with AD dementia), we conducted sensitivity analyses adjusting for cerebrospinal fluid (CSF) Aβ_1-42_ and p-tau_181_ instead of [^18^F]AZD4694 Aβ PET and [^18^F]MK6240 tau PET, respectively. Demographics for the subgroup of participants are presented in **Table S2**. Similarly, we found that *APOE*ε4 carriership was significantly associated with higher [^11^C]PBR28 SUVR in Braak I-II regions independently of CSF Aβ_1-42_ and p-tau_181_ (β = 0.073, 95% CI 0.005 to 0.140, *P* = 0.035; **Table S3**), which reinforces the cross-modality imaging results. Exploratory analyses conducted across the six Braak regions supported that the Aβ- and tau-independent association of *APOE*ε4 carriership with [^11^C]PBR28 uptake was mainly confined to early Braak regions, either assessing AD hallmark proteins with imaging ([^18^F]AZD4694 Aβ PET and [^18^F]MK6240 tau PET; **Table S4**) or fluid biomarkers (CSF Aβ_1-42_ and p-tau_181_; **Table S5**).

**Table 2.**
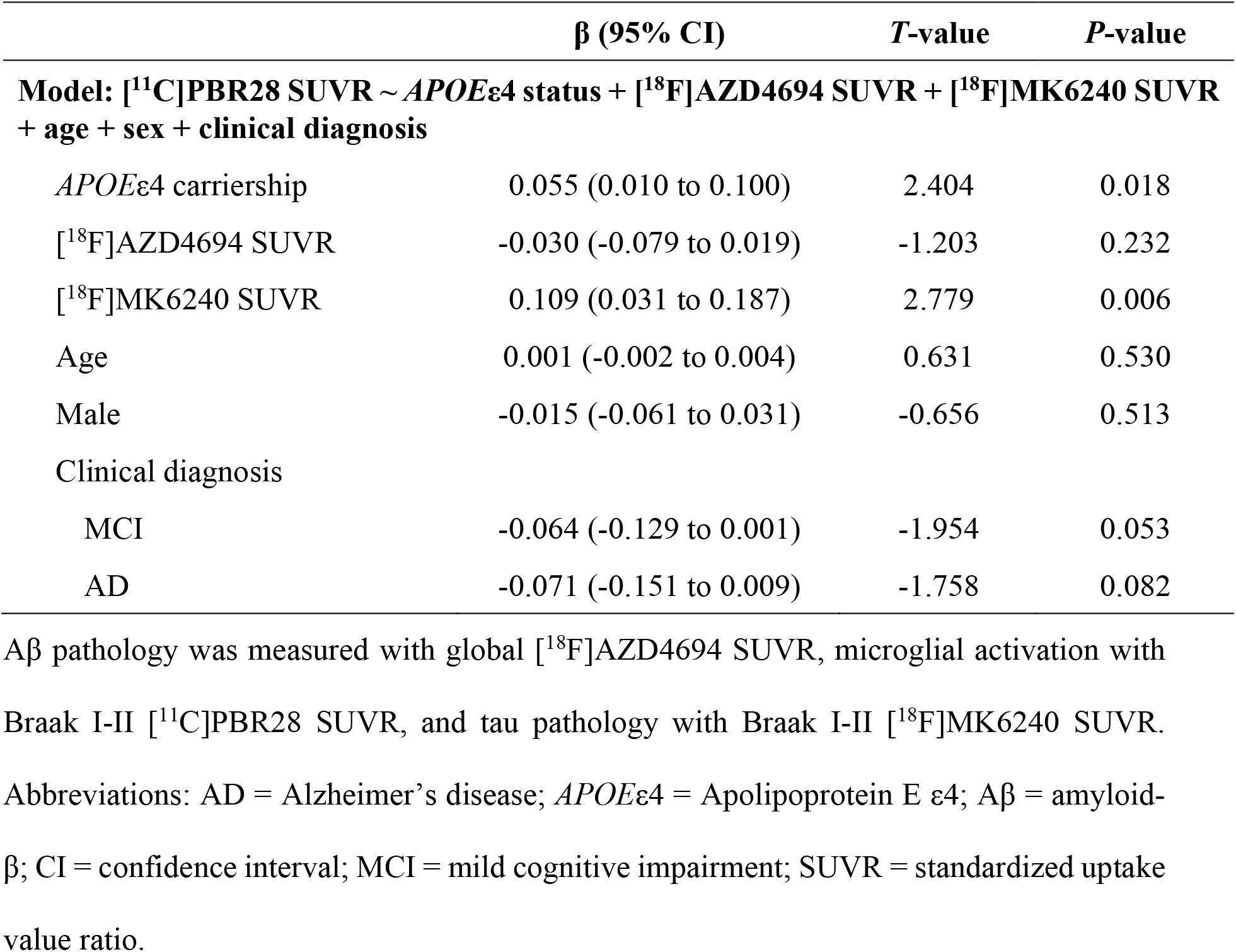
The association between *APOE*ε4 carriership and microglial activation is independent of Aβ and tau PET.

### *APOE* gene expression resembles *APOE*ε4-related microglial activation patterns

We studied the topographical distribution of *APOE* messenger RNA (mRNA) in the *postmortem* brain of six CU individuals from the Allen Human Brain Atlas. We observed different *APOE* gene expression levels across Braak regions, which were progressively lower from Braak I to VI (**Fig. 2A**), suggesting that the cerebral levels of *APOE* gene expression partially follow Braak-like stages. Additionally, linear regressions demonstrated that the regional patterns of Allen *APOE* mRNA expression across Braak regions predicted the topography and magnitude of *APOE*ε4 effects on [^11^C]PBR28 uptake observed in our population (**Fig. 2B, C**).

**Fig. 2.**
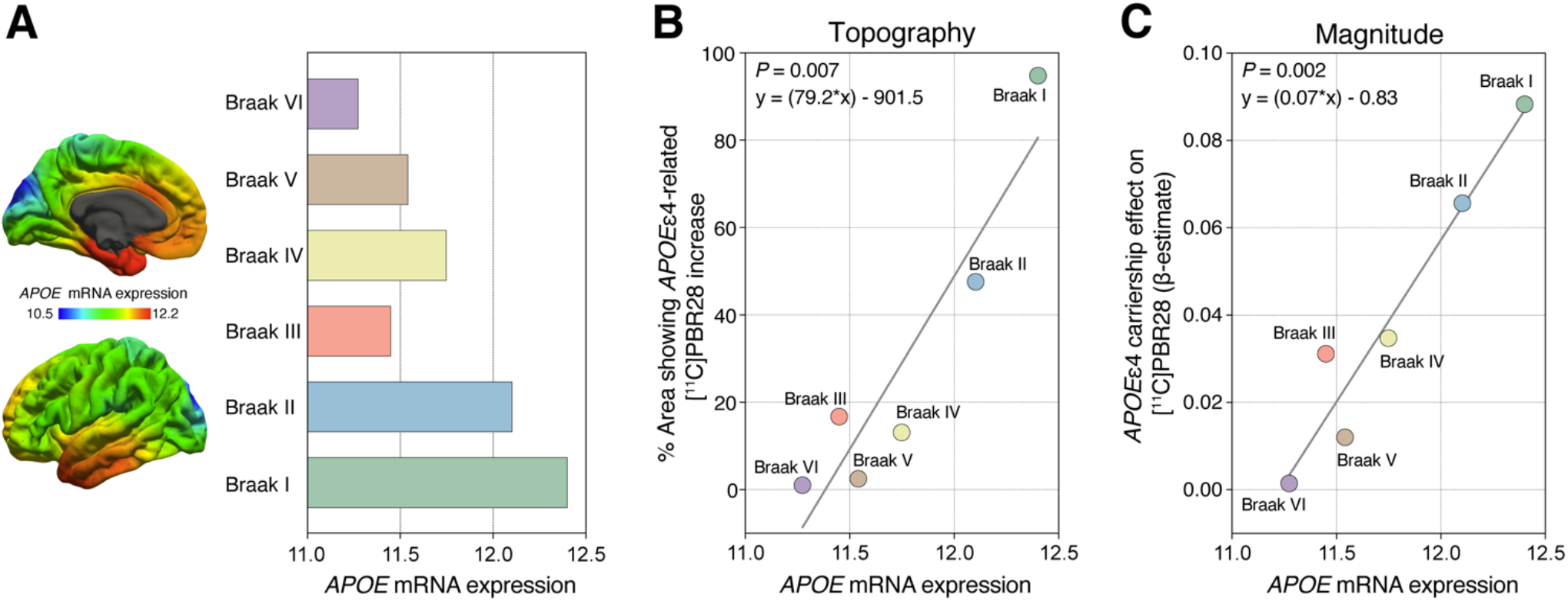
The brain levels of *APOE* gene expression predict *APOE*ε4-related [^11^C]PBR28 SUVR increase. (**A**) Brain map of the topographical distribution of *APOE* mRNA expression in 6 CU individuals obtained from the Allen Human Brain Atlas (left). Average intensity values of *APOE* mRNA expression in each Braak region (right). (**B**) Regression analysis testing whether Allen *APOE* gene expression patterns predict the percentage of the area showing *APOE*ε4-related [^11^C]PBR28 SUVR increase across Braak regions in our population. (**C**) Regression analysis testing whether the Allen brain *APOE* gene expression patterns predict the magnitude/strength of the association between *APOE*ε4 and [^11^C]PBR28 uptake across Braak regions in our population.

### Microglial activation mediates the Aβ-independent effects of *APOE*ε4 on downstream AD markers

We next used structural equation modeling to investigate the associations between *APOE*ε4, microglial activation, Aβ, tau, hippocampal volume, and clinical function (as measured with the clinical dementia rating scale sum of boxes [CDR-SB] score). We found that an increase in [^11^C]PBR28 microglial activation uptake partially mediated the effects of *APOE*ε4 carriership on higher tau PET uptake in Braak I-II regions independently of Aβ PET burden. The model also showed the well-reported effects of *APOE*ε4 on higher tau PET uptake through higher Aβ PET load, which were independent of microglial activation. Notably, both Aβ-independent and -dependent pathways leading to medial temporal tau pathology were further associated with lower hippocampal volume and, ultimately, higher severity of clinical impairment (**Fig. 3**). This model fit the data well (*n* = 118, *X*^2^ = 7.141, degrees of freedom = 5, *P* = 0.210, root mean squared error of approximation [RMSEA] = 0.060, standardized root mean square residual [SRMR] = 0.023, comparative fit index [CFI] = 0.991). Complete model coefficients and associated statistics are described in **Table S6**.

**Fig. 3.**
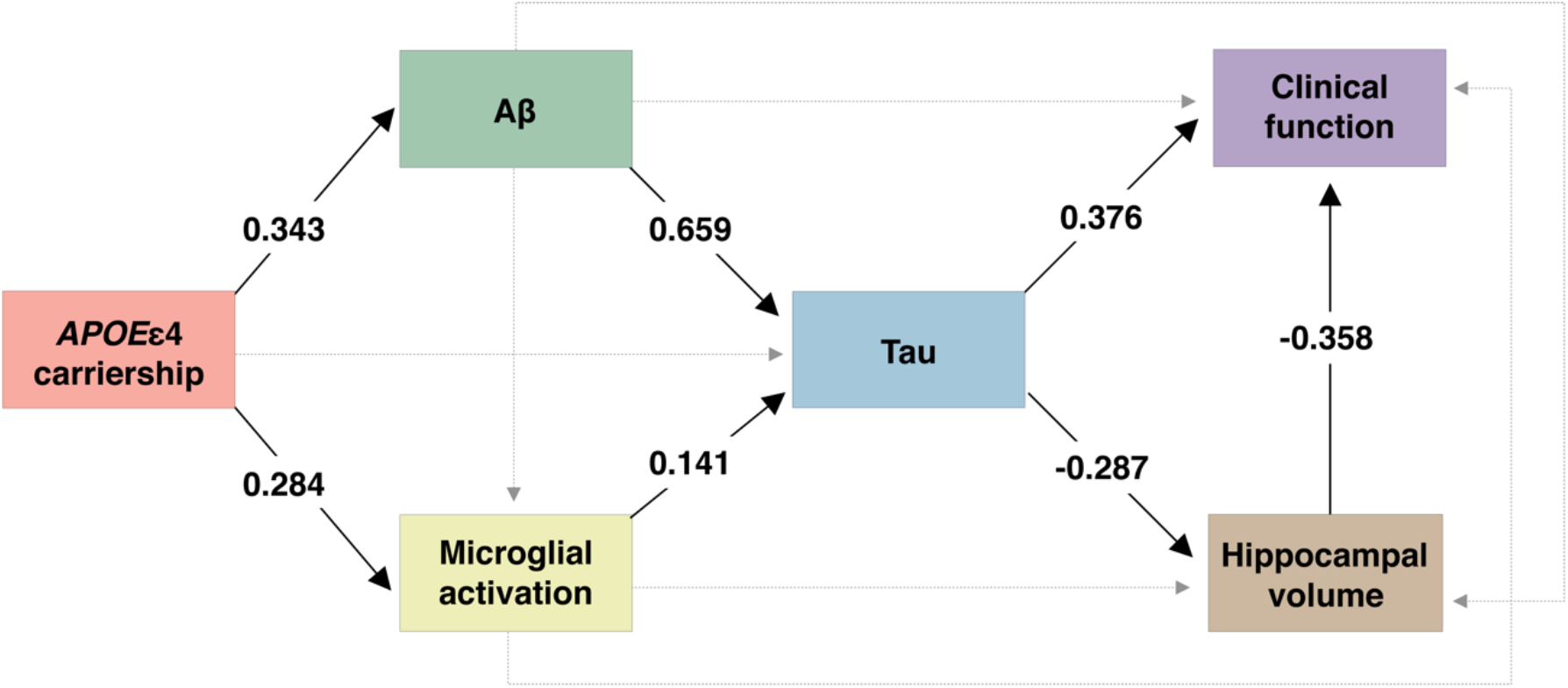
*APOE*ε4 contributes to AD progression independently of Aβ by activating microglia. The values presented in the figure are structural equation model β estimates testing the associations between *APOE*ε4 status, microglial activation PET, Aβ PET, tau PET, hippocampal volume, and clinical function. Given that the β estimates presented in the figure are standardized, the effects can be directly compared. Solid lines represent significant associations, whereas dashed lines represent non-significant effects. All associations were adjusted for age and sex. Associations involving hippocampal volume and clinical function were also adjusted for years of education. Aβ pathology was measured with global [^18^F]AZD4694 SUVR, microglial activation with Braak I-II [^11^C]PBR28 SUVR, and tau pathology with Braak I-II [^18^F]MK6240 SUVR. Clinical function was assessed with the CDR-SB score.

## Discussion

In the present study, we observed that *APOE*ε4 carriership was associated with microglial activation in early Braak-stage regions independently of AD hallmark proteinopathies. We also found that the brain distribution of *APOE* gene expression predicted the pattern of *APOE*ε4-related microglial activation observed in our study population. Lastly, we demonstrated that microglial activation partially mediated the *APOE*ε4 effects on regional tau accumulation through an Aβ-independent pathway, which was further associated with neurodegeneration and clinical impairment. Taken together, our findings support the hypothesis that *APOE*ε4 contributes to the early progression of AD via increased neuroinflammation.

*APOE*ε4 carriership was associated with higher levels of microglial activation in living humans across the aging and AD continuum. Several recent investigations in animal models of AD support our findings. For example, the *APOE* genotype modulates microglial response in AD, with *APOE*ε4 being associated with multiple microglial-related detrimental downstream effects (*e*.*g*., protein aggregation, neurodegeneration, and dysfunctional immunometabolic response) *(25, 27-30)*. Further experiments showed that the *APOE*ε4 genotype associates with changes in the transcriptional profile of microglia from a homeostatic state to a disease-associated state across AD progression *(31, 32)* and that the activation of microglial-related proteins (*e*.*g*., triggering receptor expressed on myeloid cells 2 (TREM2) *(33-36)*) is directly associated with ApoE signaling *(37)*. This evidence raises the possibility that the mechanisms explaining the link between ApoE and microglial activation occur at the transcriptional level by the expression of microglia-specific genes such as *TREM2*. Altogether, the aforementioned findings suggest that the *APOE*ε4 genotype is associated with a disruption in microglia homeostasis in AD, promoting disease-associated microglia that plays a role in the development of the disease.

Our results showed an effect of *APOE*ε4 on medial temporal microglial activation leading to AD progression that is partially independent of Aβ and tau. Previous observations indicate that microglial activation may precede and drive tau propagation *(5-7, 38)*. Moreover, an investigation using the [^11^C]PK11195 PET tracer found that microglial activation in the temporal lobe is associated with longitudinal cognitive decline more strongly than the [^18^F]AV1451 tau PET tracer binding in patients presenting AD pathophysiology *(12)*. We complemented these reports by demonstrating an Aβ-independent effect of *APOE*ε4 on early microglial activation in medial temporal structures, which in turn mediate tau accumulation and AD progression. These results resonate with recent CSF biomarker evidence of neuroinflammation in adult *APOE*ε4 carriers who have not developed Aβ pathology yet *(39)*. Our findings are also in line with animal model studies showing that the *APOE* genotype affects tau pathology as well as tau-mediated neurodegeneration independently of Aβ, with the *APOE*ε4 isoform having detrimental effects on both outcomes *(25, 38, 40-42)*. In humans, *APOE*ε4 carriership has been associated with medial temporal atrophy *(43-46)*. Additionally, a recent study revealed that *APOE*ε4 carriership is associated with tau PET uptake in the medial temporal lobe independently of Aβ, raising discussions about the importance of elucidating the biological underpinnings of this association *(47)*. We built on these previous investigations suggesting that microglial activation is the mediator of the Aβ-independent effects of *APOE*ε4 on medial temporal tau deposition and brain atrophy, leading to dementia. Together, these results suggest that disease-modifying therapies targeting the interplay between ApoE and microglial activation have the potential to slow downstream AD progression.

Interestingly, we observed that cerebral *APOE* mRNA expression was more prominent in medial temporal structures and its levels hierarchically followed the Braak staging scheme. Even though the Allen Human Brain Atlas is derived from younger adults without dementia, the *APOE* gene expression pattern was able to predict the topography and magnitude of the *APOE*ε4-related [^11^C]PBR28 uptake increase in our cohort. It is well-established that tau neurofibrillary tangles follow a stereotypical pattern of progression known as Braak stages, with tau tangles deposition starting in the medial temporal cortex *(8-11)*. Microglial activation precedes and drives tau spread from the medial temporal lobe to the neocortex in a Braak-like pattern in AD models *(5-7)*, although the mechanism associated with triggering microglial activation in medial temporal structures was not fully understood. Here, we first showed that *APOE*ε4 plays a role in triggering microglia activation in early Braak regions, and second that a Braak-like pattern of *APOE* gene expression could shed light on the entire hierarchical progression of tau across these stages. These results indicate that microglia-mediated tau propagation in AD might be explained at least partially by the cerebral expression levels of ApoE4.

Methodological strengths of the present work include the assessment of a well-characterized cohort with multiple PET radiotracers acquired on the same scanner, which allows high-quality topographical characterization of microglial activation, Aβ deposition, and tau accumulation using the best currently available technologies. Moreover, we screened a large sample of 606 individuals for the rs6971 polymorphism in the *TSPO* gene, and, consequently, we were able to include only high-affinity binders for the [^11^C]PBR28 radiotracer, which increases the signal-to-noise ratio of the tracer and the reliability of our results. Methodological limitations need to be acknowledged and considered to interpret our results. The [^11^C]PBR28 radiotracer binds to the TSPO, which is a protein mainly expressed in the mitochondrial outer membrane of activated microglia *(3)*. Thus, [^11^C]PBR28 PET is considered an imaging biomarker of a general cerebral microglial activation state *(7, 48, 49)*. However, it is recognized that microglia may acquire diverse phenotypes across disease progression *(3)*, and this heterogeneity cannot be captured using the available human brain imaging technologies. Furthermore, it is possible that other cell types (*e*.*g*., astrocytes) also play a minor role in the TSPO PET signal *(3, 50-53)*. Because CSF measures have been suggested to detect Aβ and tau accumulation earlier than PET *(48, 54, 55)*, the fact we observed similar results using imaging ([^18^F]AZD4694 and [^18^F]MK6240) and fluid (CSF Aβ_1-42_ and p-tau_181_) biomarkers support the robustness of our findings. However, we cannot exclude a possible contribution of pre-fibrillary Aβ and tau aggregates to our results. Future studies using multiple longitudinal measures are needed to better evaluate the sequential relationship between neuroimaging biomarkers. Lastly, individuals included in our investigation were volunteers motivated to participate in a study about AD, which can be a source of self-selection bias.

In conclusion, our results support the existence of an Aβ-independent effect of *APOE*ε4 on AD progression through microglial activation, leading to tau accumulation, neurodegeneration, and eventually clinical impairment.

## Materials and methods

### Experimental design

The main objective of the present study was to test the association between *APOE*ε4 carriership and brain levels of microglial activation. We hypothesized that *APOE*ε4 is associated with microglial activation in early Braak regions independently of Aβ and tau pathologies. Furthermore, we aimed to assess whether microglial activation mediates the effects of *APOE*ε4 on downstream AD progression. Participants from the community or outpatients at the McGill University Research Centre for Studies in Aging were enrolled in the Translational Biomarkers in Aging and Dementia (TRIAD) study (https://triad.tnl-mcgill.com). Participants were required to have adequate visual and auditory capacities for neuropsychological assessment, as well as the ability to speak English or French. Additionally, individuals were not enrolled if they had active substance abuse, major surgery, recent head trauma, neuroimaging contraindication, simultaneously being enrolled in other studies, and untreated neurological, psychiatric, or systemic conditions. This study was approved by the Douglas Mental Health University Institute Research Ethics Board and the Montreal Neurological Institute PET working committee, and all participants provided written informed consent.

### Participants

Out of the 606 individuals screened for the rs6971polymorphism in the *TSPO* gene, we studied 118 individuals with high-affinity binding aged 52 to 87 years (79 CU, 23 with MCI, and 16 with AD dementia). At the same time point, all participants had PET for Aβ ([^18^F]AZD4694), tau tangles ([^18^F]MK6240), and microglial activation ([^11^C]PBR28), as well as MRI and *APOE* genotyping. Two individuals that had [^11^C]PBR28 or [^18^F]MK6240 SUVR values three standard deviations (SD) above the mean of the population were considered outliers as defined *a priori* and excluded from the analyses. A flowchart describing the selection of study participants is reported in **Fig. S1**. Participants underwent detailed neuropsychological assessments, including Mini-Mental State Examination (MMSE) and Clinical Dementia Rating (CDR). CU subjects had no objective cognitive impairment and a global CDR score of 0. MCI patients had subjective and objective cognitive impairment, preserved activities of daily living, and a global CDR score of 0.5 *(56)*. Mild-to-moderate AD dementia patients had a global CDR score between 0.5 and 2 and met the National Institute on Aging and the Alzheimer’s Association (NIA-AA) criteria for probable AD *(57)*. We analyzed all the individuals with complete data, and no power analysis was performed before the study. It is worth noting that the sample size of the present work is similar to the largest previous TSPO PET studies across the AD spectrum *(7, 58-60)*.

### Genetic data

[^11^C]PBR28 binding affinity is influenced by a common polymorphism (rs6971) in the *TSPO* gene (https://www.ncbi.nlm.nih.gov/snp/rs6971). In order to increase the reliability of our results, we genotyped 606 participants for the rs6971 polymorphism before imaging, and we only included high-affinity binders *(7)*. Noteworthy, this polymorphism is a methodological caveat and does not affect TSPO levels, glial activity, or AD pathological changes *(53)*. Moreover, subjects were genotyped for the *APOE* gene using the polymerase chain reaction amplification technique, followed by restriction enzyme digestion, standard gel resolution, and visualization processes *(61)*.

### Brain imaging

T_1_-weighted MRIs were acquired at the Montreal Neurological Institute (MNI) using a 3T Siemens Magnetom. We used the magnetization prepared rapid acquisition gradient echo (MPRAGE) MRI (TR: 2300 ms, TE: 2.96ms) sequence to obtain high-resolution structural images of the whole brain (9° flip angle, coronal orientation perpendicular to the double spin echo sequence, 1×1 mm^2^ in-plane resolution of 1 mm slab thickness) *(62)*. Aβ PET with [^18^F]AZD4694 (40–70 min post-injection), tau PET with [^18^F]MK-6240 (90–110 min post-injection), and microglial activation TSPO PET with [^11^C]PBR28 (60–90 min post-injection) were acquired at the MNI using a Siemens High Resolution Research Tomograph. PET scans were reconstructed using the ordered subset expectation maximization (OSEM) algorithm on a four-dimensional (4D) volume with three frames (3 × 600 seconds) for [^18^F]AZD4694 PET *(63)*, four frames (4 × 300 seconds) for [^18^F]MK-6240 PET *(63)*, and six frames (6 × 300 s) for [^11^C]PBR28 PET *(7)*. Then, attenuation correction was performed using a 6-minute transmission scan with a rotating ^137^Cs point source. Furthermore, PET images were corrected for motion, dead time, decay, and scattered and random coincidences. Following an in-house pipeline, T_1_-weighted MRIs were corrected for non-uniformity and field distortion. Subsequently, linear co-registration and non-linear spatial normalization for the ADNI template were performed through linear and non-linear transformation in two main steps: (i) PET registration to the correspondent T_1_-weighted MRI and (ii) T_1_-weighted MRI registration to the ADNI reference space. PET images were spatially smoothed to achieve a final resolution of 8 mm full width at half-maximum. SUVRs were calculated using the whole cerebellum gray matter for [^18^F]AZD4694 Aβ PET *(64)* and [^11^C]PBR28 microglial activation PET *(7)*, and the inferior cerebellum gray matter for [^18^F]MK-6240 tau PET *(65)*. The Desikan-Killiany-Tourville atlas was used to determine the regions of interest (ROIs) *(66)*. A global Aβ PET SUVR was estimated from the precuneus, prefrontal, orbitofrontal, parietal, temporal, and cingulate cortices *(67)*. Based on *postmortem* observations *(8, 9)* and PET studies *(65, 68)*, PET Braak-like stages were calculated from brain regions corresponding to the Braak stages of tau neurofibrillary tangle accumulation: Braak I (transentorhinal), Braak II (entorhinal and hippocampus), Braak III (amygdala, parahippocampal gyrus, fusiform gyrus, lingual gyrus), Braak IV (insula, inferior temporal, lateral temporal, posterior cingulate, and inferior parietal), Braak V (orbitofrontal, superior temporal, inferior frontal, cuneus, anterior cingulate, supramarginal gyrus, lateral occipital, precuneus, superior parietal, superior frontal, rostro medial frontal), and Braak VI (paracentral, postcentral, precentral, and pericalcarine) *(65, 69)*. A representation of the Braak-like regions used in our analysis is shown in **Fig. S3**. Hippocampal volume was adjusted for total intracranial volume using MRI data from CU subjects *(70)*.

The Allen Human Brain Atlas (http://www.brain-map.org) *(71)* was used to obtain information regarding *APOE* gene expression in the brain. In brief, microarray was used to calculate mRNA expression intensity values on 3702 samples from 6 healthy human *postmortem* brains (4 males, mean age = 42.5 (13.4) years, *postmortem* delay = 20.6 (7) hours). The *APOE* mRNA brain expression maps were derived from a Gaussian process *(72)* and downloaded from www.meduniwien.ac.at/neuroimaging/mRNA.html.

### CSF measurements

A subgroup of 51 individuals had CSF Aβ_1-42_ and p-tau_181_ quantified using the LUMIPULSE G1200 instrument (Fujirebio) at the Clinical Neurochemistry Laboratory, University of Gothenburg, Mölndal, Sweden *(73)*.

### Statistical analysis

Analyses were performed in the R software (version 4.0.2, http://www.r-project.org/). Neuroimaging analyses were conducted using RMINC *(74)*, an imaging package that allows the integration of voxel-based statistics into the R statistical environment. Voxel-wise and ROI-based linear regressions tested the association between *APOE*ε4 status (noncarrier or carrier) and microglial activation indexed by the [^11^C]PBR28 SUVR adjusting for age, sex, and clinical diagnosis (CU, MCI, or AD). To further investigate whether this association was independent of AD hallmark proteins, ROI-based models were also adjusted for Aβ ([^18^F]AZD4694 PET or CSF Aβ_1-42_) and tau ([^18^F]MK-6240 PET or CSF p-tau_181_) biomarkers. Multiple comparison correction at *P* < 0.05 was performed using random field theory for voxel-wise analysis and Bonferroni method for ROI-based analysis when appropriate. We calculated the percentage of voxels in each Braak region showing an association (*T*-value > 2) between *APOE*ε4 carriership and [^11^C]PBR28 SUVR in the aforementioned voxel-wise analysis. Regression analysis tested whether the *APOE* mRNA expression intensity predicted the topography and magnitude of *APOE*ε4-related [^11^C]PBR28 SUVR increase across Braak regions. Structural equation modeling, R package “lavaan” *(75)*, was used to test the associations between *APOE*ε4 status, microglial activation, Aβ, tau, hippocampal volume, and clinical function (as measured with the CDR-SB score). All the associations in the model were adjusted for age and sex; associations involving hippocampal volume and clinical deterioration were further adjusted for years of education. Structural equation model was judged as having a good fit as follows: CFI > 0.97 (acceptable: 0.95 - 0.97); RMSEA < 0.05 (acceptable 0.05 - 0.08); SRMR < 0.05 (acceptable: 0.05 - 0.10) *(76, 77)*. Statistical significance of parameters estimates from the structural equation model was tested using bootstrapping with 1000 permutations. For all analyses, two-tailed *P*-values < 0.05 were considered statistically significant.

## Data Availability

The data used in the present work is not publicly available as the information could compromise the participants′ privacy. Therefore, the data from the TRIAD study will be made available from the senior authors upon reasonable request, and such arrangements are subject to standard data-sharing agreements.

## List of Supplementary Materials

Fig. S1. Flowchart of included participants.

Fig. S2. Sensitivity analysis excluding *APOE*ε2 carriers.

Fig. S3. Braak regions mask used in the analyses.

Table S1. Prevalence of *APOE* genotypes.

Table S2. Demographics of the subsample with available CSF biomarkers.

Table S3. Sensitivity analysis testing the association of *APOE*ε4 carriership with microglial activation adjusting for CSF Aβ_1-42_ and p-tau_181_.

Table S4. Associations between microglial activation and *APOE*ε4 carriership across all Braak regions adjusting for global [^18^F]AZD4694 Aβ PET and local [^18^F]MK6240 tau PET.

Table S5. Associations between microglial activation and *APOE*ε4 carriership across all Braak regions adjusting for CSF Aβ_1-42_ and p-tau_181_.

Table S6. Structural equation model coefficients and associated statistics for Fig. 3.

## Acknowledgments

We acknowledge all study participants and the staff of the McGill University Research Center for Studies in Aging. We thank Dean Jolly, Alexey Kostikov, Monica Samoila-Lactatus, Karen Ross, Mehdi Boudjemeline, and Sandy Li for assisting in the radiochemistry production, as well as Richard Strauss, Edith Strauss, Guylaine Gagné, Carley Mayhew, Tasha Vinet-Celluci, Karen Wan, Sarah Sbeiti, Meong Jin Joung, Miloudza Olmand, Rim Nazar, Hung-Hsin Hsiao, Reda Bouhachi, and Arturo Aliaga for helping with the acquisition of the data. We also acknowledge Simon Jones and Leonidas Chouliaras at the University of Cambridge for discussing the findings. The views expressed are those of the authors and not necessarily those of the NIHR or the Department of Health and Social Care.

## Funding

Alzheimer’s Association (#NIRG-12-92090 and #NIRP-12-259245; PR-N)

Alzheimer’s Association (#AACSF-20-648075; TAP)

Brain & Behavioral Research Foundation (#29486; DTL)

Brain Canada Foundation (CFI Project 34874; 33397; PR-N)

Canadian Consortium of Neurodegeneration and Aging (#MOP-11-51-31 - team 1; PR-N)

Canadian Institutes of Health Research (#MOP-11-51-31; RFN 152985, 159815, 162303; PR-N)

CNPq (200691/2021-0; JPF-S)

CNPq (166407/2020-8; DTL)

Fonds de Recherche du Québec – Santé (Chercheur Boursier, #2020-VICO-279314; PR-N)

Fonds de Recherche du Québec – Santé (FZL)

National Institutes of Health (#R01AG075336 and #R01AG073267; TAP)

Race Against Dementia Alzheimer’s Research UK (#ARUK-RADF2021A-010; MM)

Weston Brain Institute (PR-N)

## Author contributions

Conceptualization: JPF-S, FZL, PR-N, and TAP

Methodology: JPF-S, FZL, and TAP

Formal analysis: JPF-S, FZL, CT, and DLT

Investigation: JPF-S, FZL, DTL, JT, CT, BB, PCLF, MM, Y-TW, GP, ALB, NJA, MC, SS, GB, MSK, JS, NR, VP, NMP, TKK, ERZ, PR-N, TAP

Resources: HZ, KB, PR-N

Writing - Original Draft: JPF-S

Writing - Review & Editing: JPF-S, FZL, DTL, JT, CT, BB, PCLF, MM, GP, ALB, NJA, JTO, JBR, AC, OLL, DLT, TKK, WEK, VLV, J-PS, SG, DOS, HZ, KB, ERZ, PR-N, and TAP

Supervision: TAP

Project administration: JPF-S, PR-N, TAP

Funding acquisition: JPF-S, PR-N, TAP

## Competing interests

SG has served as a scientific advisor to Cerveau Therapeutics. HZ has served at scientific advisory boards and/or as a consultant for Abbvie, Alector, Annexon, Apellis, Artery Therapeutics, AZTherapies, CogRx, Denali, Eisai, Nervgen, Novo Nordisk, Pinteon Therapeutics, Red Abbey Labs, reMYND, Passage Bio, Roche, Samumed, Siemens Healthineers, Triplet Therapeutics, and Wave, has given lectures in symposia sponsored by Cellectricon, Fujirebio, Alzecure, Biogen, and Roche, and is a co-founder of Brain Biomarker Solutions in Gothenburg AB (BBS), which is a part of the GU Ventures Incubator Program. KB has served as a consultant, at advisory boards, or at data monitoring committees for Abcam, Axon, BioArctic, Biogen, JOMDD/Shimadzu. Julius Clinical, Lilly, MagQu, Novartis, Prothena, Roche Diagnostics, and Siemens Healthineers, and is a co-founder of Brain Biomarker Solutions in Gothenburg AB (BBS), which is a part of the GU Ventures Incubator Program. ERZ serves in the scientific advisory board of Next Innovative Therapeutics. All other authors declare that they have no competing interests.

## Data and materials availability

The data used in the present work is not publicly available as the information could compromise the participants’ privacy. Therefore, the data from the TRIAD study will be made available from the senior authors upon reasonable request, and such arrangements are subject to standard data-sharing agreements.

## Notes

### Funding Statement

The Funding Statement is reported in the manuscript.

### Author Declarations

This study was approved by the Douglas Mental Health University Institute Research Ethics Board and the Montreal Neurological Institute PET working committee, and all participants provided written informed consent.

