## Supplementary Information for "*APOE*ε4 carriership associates with microglial activation independently of Aβ plaques and tau tangles"

**Fig. S1.** Flowchart of included participants.

**Fig. S2.** Sensitivity analysis excluding *APOE*ε2 carriers.

**Fig. S3.** Braak regions mask used in the analyses.

**Table S1.** Prevalence of *APOE* genotypes.

**Table S2.** Demographics of the subsample with available CSF biomarkers.

**Table S3.** Sensitivity analysis testing the association of *APOE*ε4 carriership with microglial activation adjusting for CSF $A\beta_{1-42}$ and p-$tau_{181}$.

**Table S4.** Associations between microglial activation and *APOE*ε4 carriership across all Braak regions adjusting for global [$^{18}$F]AZD4694 Aβ PET and local [$^{18}$F]MK6240 tau PET.

**Table S5.** Associations between microglial activation and *APOE*ε4 carriership across all Braak regions adjusting for CSF $A\beta_{1-42}$ and p-$tau_{181}$.

**Table S6.** Structural equation model coefficients and associated statistics for Fig. 3.

Abbreviations: AD = Alzheimer's disease; *APOE* ε4 = Apolipoprotein E ε4; Aβ = amyloid-β; CDR-SB = clinical dementia rating scale sum of boxes; CFI = comparative fit index; CI = confidence interval; CSF = cerebrospinal fluid; CU = cognitively unimpaired; MCI = mild cognitive impairment; MRI = magnetic resonance imaging; PET = positron emission tomography; p-tau$_{181}$ = tau phosphorylated at threonine 181; QC = quality control; RMSEA = root mean squared error of approximation; ROI = region of interest; SE = standard error; SRMR = standardized root mean square residual; SUVR = standardized uptake value ratio; TSPO = translocator protein.

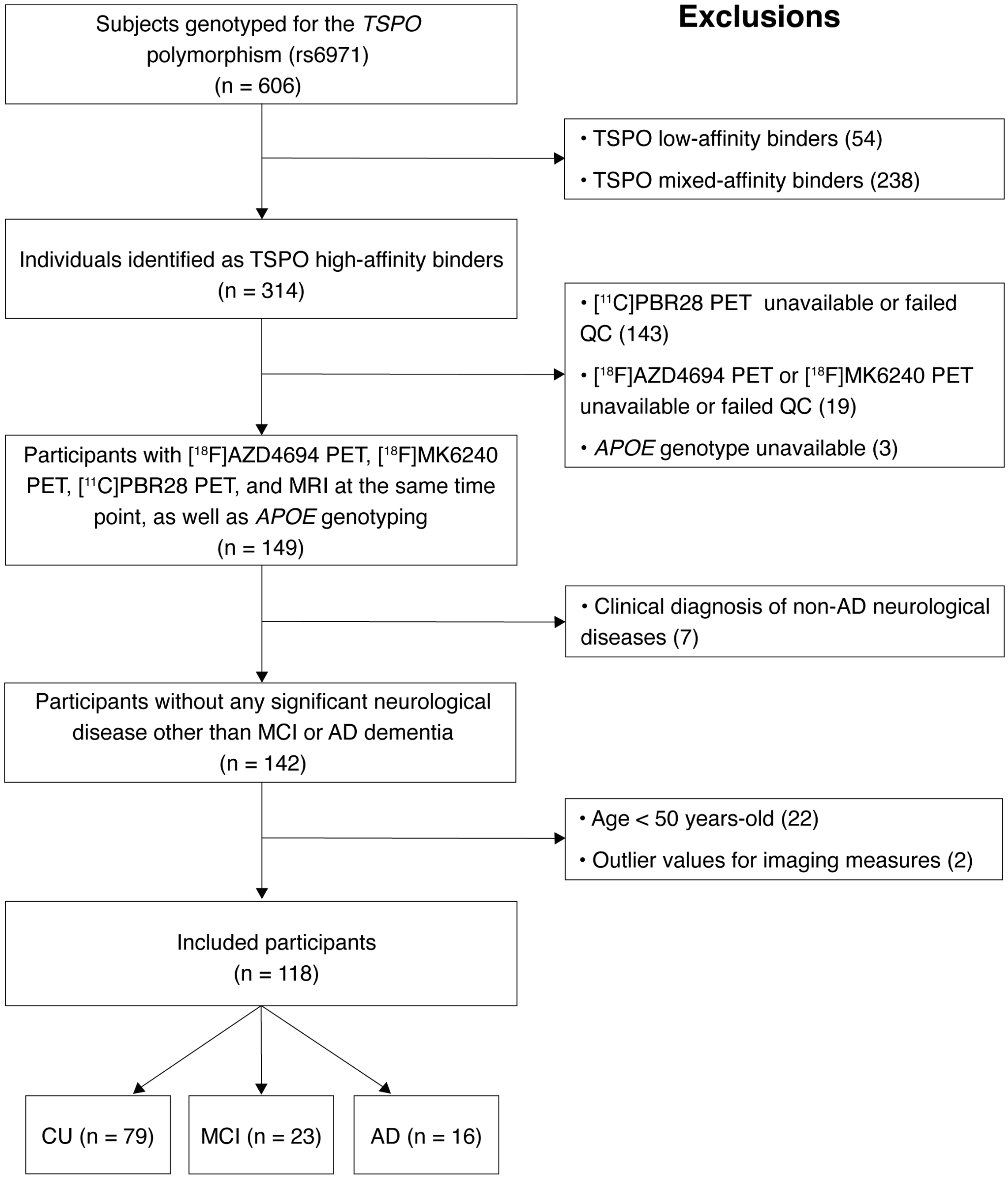


**Fig. S1. Flowchart of included participants.**

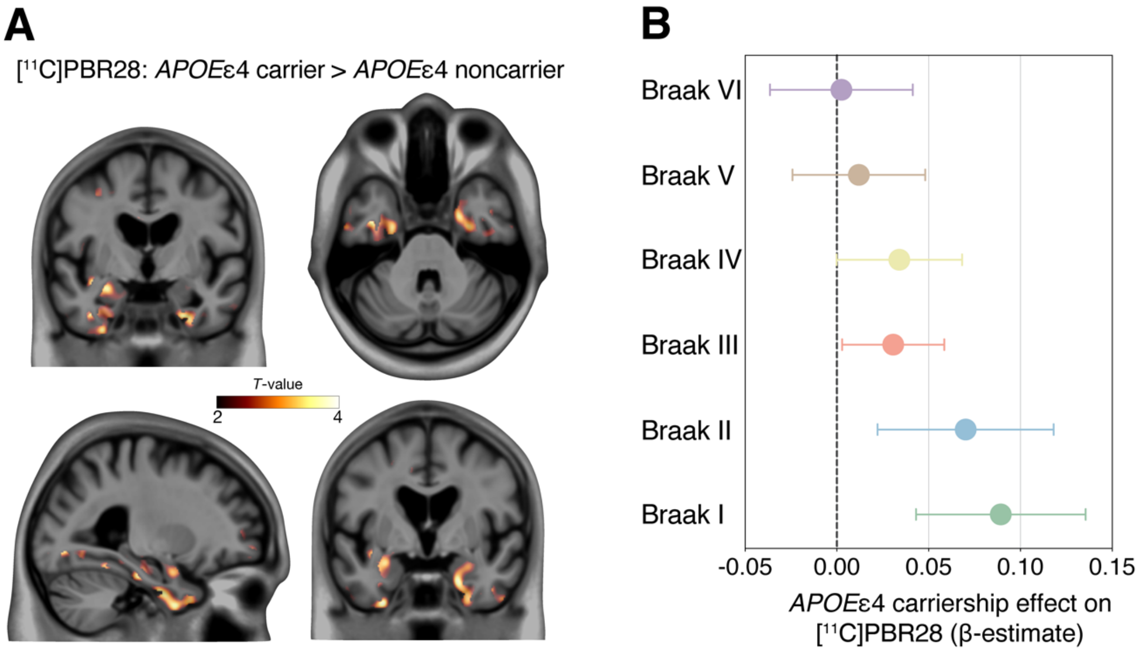


**Fig. S2. Sensitivity analysis excluding *APOE*ε2 carriers.** (**A**) *T*-map shows the result of voxel-wise linear regression testing the association of *APOE*ε4 carriage status (noncarrier or carrier) with [$^{11}$C]PBR28 SUVR accounting for age, sex, and clinical diagnosis (CU, MCI, or AD). Results survived random field theory correction for multiple comparisons at $P < 0.05$. (**B**) β estimates with 95% CI from ROI-based linear regressions represent the strength of the regional association between *APOE*ε4 status and [$^{11}$C]PBR28 SUVR across Braak regions. Models were adjusted for age, sex, and clinical diagnosis (CU, MCI, or AD). Voxel- and ROI-based sensitivity analyses were conducted after the removal of 14 individuals bearing the ε2 allele of the *APOE* gene.

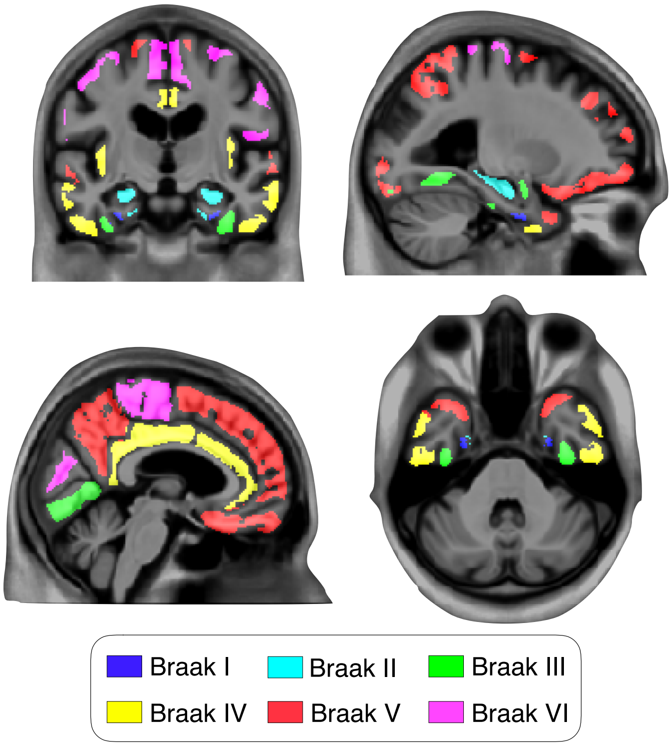


**Fig. S3. Braak regions mask used in the analyses.** Representation of the Braak-like stages ROIs overlaid on a structural MRI template. According to postmortem evidence (*8, 9*), we used the following brain regions corresponding to the Braak stages of tau neurofibrillary tangle accumulation: Braak I (transentorhinal), Braak II (entorhinal and hippocampus), Braak III (amygdala, parahippocampal gyrus, fusiform gyrus, lingual gyrus), Braak IV (insula, inferior temporal, lateral temporal, posterior cingulate, and inferior parietal), Braak V (orbitofrontal, superior temporal, inferior frontal, cuneus, anterior cingulate, supramarginal gyrus, lateral occipital, precuneus, superior parietal, superior frontal, rostro medial frontal), and Braak VI (paracentral, postcentral, precentral, and pericalcarine). A detailed description of how Braak-like stages ROIs were determined has already been reported elsewhere (*65, 69*).

**Table S1. Prevalence of *APOE* genotypes.**

| *APOE genotype* | No. (%) |
|---|---|
| ε1/ε2 | 1 (0.8) |
| ε2/ε3 | 13 (11.0) |
| ε3/ε3 | 59 (50.0) |
| ε3/ε4 | 38 (32.2) |
| ε4/ε4 | 7 (5.9) |

*APOE* genotypes that were not observed in our study population are not displayed in the table (*e.g.*, ε2/ε4).

**Table S2. Demographics of the subsample with available CSF biomarkers.**

| | CU | MCI | AD |
|---|---|---|---|
| No. | 31 | 12 | 8 |
| Age, years | 72.9 (5.7) | 71.3 (4.9) | 66.0 (10.1) |
| Male, No. (%) | 8 (25.8) | 7 (58.3) | 4 (50.0) |
| Education, years | 14.2 (3.7) | 15.3 (3.2) | 15.3 (2.8) |
| *APOE*ε4 carrier, No. (%) | 9 (29.0) | 10 (83.3) | 5 (62.5) |
| MMSE score | 29.0 (1.1) | 28.2 (1.6) | 22.5 (5.0) |
| CDR-SB score | 0.1 (0.2) | 1.4 (0.7) | 4.6 (1.7) |
| CSF $A\beta_{1-42}$, pg/mL | 943.3 (423.0) | 590.5 (230.6) | 538.5 (147.9) |
| CSF p-$tau_{181}$, pg/mL | 50.3 (35.2) | 87.3 (37.4) | 113.2 (66.2) |
| Global [$^{18}$F]AZD4694 SUVR | 1.68 (0.52) | 2.31 (0.52) | 2.42 (0.45) |
| Braak I-II [$^{18}$F]MK6240 SUVR | 0.93 (0.19) | 1.60 (0.49) | 1.62 (0.30) |
| Braak III-IV [$^{18}$F]MK6240 SUVR | 0.95 (0.10) | 1.50 (0.62) | 2.16 (1.18) |
| Braak V-VI [$^{18}$F]MK6240 SUVR | 0.98 (0.09) | 1.29 (0.37) | 2.02 (1.29) |
| Hippocampal volume, $cm^3$ | 3.48 (0.25) | 3.23 (0.32) | 3.13 (0.69) |

Continuous variables are presented as mean (SD).

**Table S3. Sensitivity analysis testing the association of *APOE*ε4 carriership with microglial activation adjusting for CSF A$\beta_{1-42}$ and p-tau$_{181}$.**

| | **β (95% CI)** | ***T*-value** | ***P*-value** |
|---|---|---|---|
| **Model: Braak I-II [$^{11}$C]PBR28 SUVR ~ *APOE*ε4 status + CSF A$\beta_{1-42}$ + CSF p-tau$_{181}$ + age + sex + clinical diagnosis** | | | |
| *APOE*ε4 carriership | 0.073 (0.005 to 0.140) | 2.179 | 0.035 |
| CSF A$\beta_{1-42}$ | 0.0001 (0.00001 to 0.0002) | 2.298 | 0.027 |
| CSF p-tau$_{181}$ | 0.0004 (-0.0003 to 0.001) | 1.110 | 0.273 |
| Age | 0.007 (0.002 to 0.012) | 2.799 | 0.008 |
| Male | -0.013 (-0.077 to 0.051) | -0.422 | 0.675 |
| Clinical diagnosis | | | |
| MCI | -0.011 (-0.099 to 0.077) | -0.252 | 0.802 |
| AD | 0.054 (-0.048 to 0.156) | 1.062 | 0.294 |

Aβ pathology was measured with CSF A$\beta_{1-42}$, tau pathology with CSF p-tau$_{181}$, and microglial activation with Braak I-II [$^{11}$C]PBR28 SUVR.

**Table S4. Associations between microglial activation and *APOE*ε4 carriership across all Braak regions adjusting for global [$^{18}$F]AZD4694 Aβ PET and local [$^{18}$F]MK6240 tau PET.**

| | β (95% CI) | *T*-value | *P*-value |
|---|---|---|---|
| **Model A: Braak I [$^{11}$C]PBR28 SUVR ~ *APOE*ε4 status + global [$^{18}$F]AZD4694 SUVR + Braak I [$^{18}$F]MK6240 SUVR + age + sex + clinical diagnosis** | | | |
| *APOE*ε4 carriership | 0.087 (0.040 to 0.135) | 3.628 | < 0.001 |
| **Model B: Braak II [$^{11}$C]PBR28 SUVR ~ *APOE*ε4 status + global [$^{18}$F]AZD4694 SUVR + Braak II [$^{18}$F]MK6240 SUVR + age + sex + clinical diagnosis** | | | |
| *APOE*ε4 carriership | 0.053 (0.006 to 0.099) | 2.252 | 0.026 |
| **Model C: Braak III [$^{11}$C]PBR28 SUVR ~ *APOE*ε4 status + global [$^{18}$F]AZD4694 SUVR + Braak III [$^{18}$F]MK6240 SUVR + age + sex + clinical diagnosis** | | | |
| *APOE*ε4 carriership | 0.029 (0.002 to 0.057) | 2.099 | 0.038 |
| **Model D: Braak IV [$^{11}$C]PBR28 SUVR ~ *APOE*ε4 status + global [$^{18}$F]AZD4694 SUVR + Braak IV [$^{18}$F]MK6240 SUVR + age + sex + clinical diagnosis** | | | |
| *APOE*ε4 carriership | 0.032 (-0.003 to 0.067) | 1.813 | 0.073 |
| **Model E: Braak V [$^{11}$C]PBR28 SUVR ~ *APOE*ε4 status + global [$^{18}$F]AZD4694 SUVR + Braak V [$^{18}$F]MK6240 SUVR + age + sex + clinical diagnosis** | | | |
| *APOE*ε4 carriership | 0.015 (-0.022 to 0.052) | 0.797 | 0.427 |
| **Model F: Braak VI [$^{11}$C]PBR28 SUVR ~ *APOE*ε4 status + global [$^{18}$F]AZD4694 SUVR + Braak VI [$^{18}$F]MK6240 SUVR + age + sex + clinical diagnosis** | | | |
| *APOE*ε4 carriership | 0.005 (-0.036 to 0.046) | 0.257 | 0.797 |

Global [$^{18}$F]AZD4694 SUVR and local [$^{18}$F]MK6240 SUVR were used to assess Aβ and tau pathologies, respectively.

**Table S5. Associations between microglial activation and *APOE*ε4 carriership across all Braak regions adjusting for CSF A$\beta_{1-42}$ and p-tau$_{181}$.**

| | β (95% CI) | *T*-value | *P*-value |
|---|---|---|---|
| **Model A: Braak I [$^{11}$C]PBR28 SUVR ~ *APOE*ε4 status + CSF A$\beta_{1-42}$ + CSF p-tau$_{181}$ + age + sex + clinical diagnosis** | | | |
| *APOE*ε4 carriership | 0.086 (0.006 to 0.165) | 2.179 | 0.035 |
| **Model B: Braak II [$^{11}$C]PBR28 SUVR ~ *APOE*ε4 status + CSF A$\beta_{1-42}$ + CSF p-tau$_{181}$ + age + sex + clinical diagnosis** | | | |
| *APOE*ε4 carriership | 0.071 (0.002 to 0.141) | 2.070 | 0.044 |
| **Model C: Braak III [$^{11}$C]PBR28 SUVR ~ *APOE*ε4 status + CSF A$\beta_{1-42}$ + CSF p-tau$_{181}$ + age + sex + clinical diagnosis** | | | |
| *APOE*ε4 carriership | 0.013 (-0.027 to 0.054) | 0.673 | 0.504 |
| **Model D: Braak IV [$^{11}$C]PBR28 SUVR ~ *APOE*ε4 status + CSF A$\beta_{1-42}$ + CSF p-tau$_{181}$ + age + sex + clinical diagnosis** | | | |
| *APOE*ε4 carriership | 0.019 (-0.038 to 0.075) | 0.670 | 0.507 |
| **Model E: Braak V [$^{11}$C]PBR28 SUVR ~ *APOE*ε4 status + CSF A$\beta_{1-42}$ + CSF p-tau$_{181}$ + age + sex + clinical diagnosis** | | | |
| *APOE*ε4 carriership | 0.015 (-0.045 to 0.076) | 0.515 | 0.609 |
| **Model F: Braak VI [$^{11}$C]PBR28 SUVR ~ *APOE*ε4 status + CSF A$\beta_{1-42}$ + CSF p-tau$_{181}$ + age + sex + clinical diagnosis** | | | |
| *APOE*ε4 carriership | 0.022 (-0.034 to 0.079) | 0.799 | 0.429 |

CSF A$\beta_{1-42}$ and CSF p-tau$_{181}$ were used to assess Aβ and tau pathologies, respectively.

**Table S6. Structural equation model coefficients and associated statistics for Fig. 3.**

| | β (SE) | Z-value | Std. β | *P*-value |
|---|---|---|---|---|
| **CDR-SB** | | | | |
| Hippocampal volume | -1.733 (0.477) | -3.630 | -0.358 | < 0.001 |
| [$^{18}$F]MK6240 SUVR | 1.882 (0.791) | 2.380 | 0.376 | 0.017 |
| [$^{11}$C]PBR28 SUVR | -2.174 (1.226) | -1.773 | -0.122 | 0.076 |
| [$^{18}$F]AZD4694 SUVR | 0.214 (0.384) | 0.558 | 0.064 | 0.577 |
| **Hippocampal volume** | | | | |
| [$^{18}$F]MK6240 SUVR | -0.296 (0.132) | -2.239 | -0.287 | 0.025 |
| [$^{11}$C]PBR28 SUVR | 0.632 (0.339) | 1.862 | 0.171 | 0.063 |
| [$^{18}$F]AZD4694 SUVR | -0.157 (0.101) | -1.563 | -0.225 | 0.118 |
| **[$^{18}$F]MK6240 SUVR** | | | | |
| *APOE*ε4 carriership | 0.075 (0.057) | 1.325 | 0.092 | 0.185 |
| [$^{11}$C]PBR28 SUVR | 0.501 (0.253) | 1.983 | 0.141 | 0.047 |
| [$^{18}$F]AZD4694 SUVR | 0.444 (0.050) | 8.840 | 0.659 | < 0.001 |
| **[$^{11}$C]PBR28 SUVR** | | | | |
| *APOE*ε4 carriership | 0.065 (0.022) | 3.017 | 0.284 | 0.003 |
| [$^{18}$F]AZD4694 SUVR | -0.013 (0.018) | -0.700 | -0.068 | 0.484 |
| **[$^{18}$F]AZD4694 SUVR** | | | | |
| *APOE*ε4 carriership | 0.418 (0.112) | 3.738 | 0.343 | < 0.001 |

Structural equation model estimates and associated statistics testing the associations between *APOE*ε4 status (noncarrier or carrier), microglial activation, Aβ, tau, hippocampal volume, and clinical function. All associations were adjusted for age and sex. Associations involving clinical function and hippocampal volume were also adjusted for years of education. Aβ pathology was measured with global [$^{18}$F]AZD4694 SUVR, tau pathology was measured with Braak I-II [$^{18}$F]MK6240 SUVR, and microglial activation was measured with Braak I-II [$^{11}$C]PBR28 SUVR. Clinical function was assessed with the CDR-SB score. The model fitted the data well ($n$ = 118, $X^2$ = 7.141, degrees of freedom = 5, $P$ = 0.210, RMSEA = 0.060, SRMR = 0.023, CFI = 0.991).
